# Implications of rapid population growth on survey design and HIV estimates in the Rakai Community Cohort Study (RCCS), Uganda

**DOI:** 10.1101/2022.09.06.22279646

**Authors:** Aleya Khalifa, Robert Ssekubugu, Justin Lessler, Maria J. Wawer, John Santelli, Susie Hoffman, Fred Nalugoda, Tom Lutalo, Anthony Ndyanbo, Joseph Ssekasanvu, Godfrey Kigozi, Joseph Kagaayi, Larry W. Chang, M. Kate Grabowski

## Abstract

**Background:** Longitudinal population-based cohorts are critical in HIV surveillance programs in Africa but continued rapid population growth poses serious challenges to maintaining cohort representativeness with limited resources. In one such cohort, we evaluated if systematic exclusion of some residents due to growing population size biases key HIV metrics like prevalence and viremia.

**Methods:** Data were obtained from the Rakai Community Cohort study (RCCS) in south central Uganda, an open population-based cohort which began excluding some residents of newly constructed household structures within its surveillance boundaries in 2008. We evaluated the extent to which changing inclusion criteria may bias recent population HIV seroprevalence and viremia estimates from the RCCS using ensemble machine learning models fit to 2019-2020 RCCS census and survey data.

**Results:** Of the 24,729 census-eligible residents, 2,920 (12%) were living within new household structures and excluded. Predicted seroprevalence for excluded residents was 11.4% (95% Confidence Interval: 10.2, 12.3) compared to 11.8% in the observed sample. However, predicted seroprevalence for younger excluded residents 15-24 years was 5.1% (3.6, 6.1), which was significantly higher than that in the observed sample for the same age group (2.6%). Over all ages, predicted prevalence of viremia in excluded residents (2.8% [2.2, 3.3]) was higher than that in the observed sample (1.7%), resulting in a somewhat higher overall population viremia estimate of 1.9% [1.8, 2.0]).

**Conclusions:** Exclusion of residents in new households may modestly bias HIV viremia estimates and some age-specific seroprevalence estimates in the RCCS. Overall HIV seroprevalence estimates were not significantly affected.

**Key messages (3-5):**

- In-migrants in the observed sample in the RCCS surveillance area differ from currently excluded in-migrants on various demographic characteristics.
- Machine learning methods may be useful tools in estimating biases introduced by the systematic exclusion of populations for which we have some data.
- In the context of rapid population growth, population-based open cohorts in sub-Saharan Africa must prioritize limited resources while ensuring HIV estimates are representative of the population.

**Funding:** Funding for this project was supported by the National Institute of Allergy and Infectious Diseases (R01AI143333 and R01AI155080) and the National Institute of Mental Health (R01MH115799). The findings and conclusions in this article are those of the authors and do not necessarily represent the official position of the funding agencies.

Research by Aleya Khalifa reported in this publication was supported by the National Institute of Allergy And Infectious Diseases (T32AI114398). Larry Chang was supported by the National Heart, Lung, and Blood Institute (R01HL152813), Fogarty International Center (D43TW010557) and the Johns Hopkins University Center for AIDS Research (P30AI094189). Susie Hoffman and John Santelli were supported by the U.S. National Institute of Child Health and Human Development (NICHD) (R01HD091003; Santelli, PI). Susie Hoffman was also supported by the National Institute of Mental Health (P30-MH43520; Remien, PI). The content is solely the responsibility of the authors and does not necessarily represent the official views of the National Institutes of Health.

**Ethics approval:** This study was approved by the Uganda National Council for Science and Technology (approval number HS 540), the Uganda Virus Research Institution Research and Ethics Committee (approval number GC/127/08/12/137), Johns Hopkins Institutional Review Board (approval number IRB-00217467), and the Columbia University Institutional Review Board (approval number IRB-AAAR5428).

## Introduction

Over the last 30 years, longitudinal population-based cohorts in sub-Saharan Africa (SSA) have provided critical surveillance data on the determinants of HIV acquisition and transmission, and evolving HIV epidemic trends.^1^ An important challenge these studies face is rapid population growth^2–4^ often within the context of fixed budgets and resource constraints.^5,6,7^ For example, the Rakai Community Cohort Study (RCCS) – established in 1994 in south-central Uganda – has seen a more than 200% increase in population size within its surveillance boundaries over the last 20 years.^8^ Most of this population increase in the RCCS catchment area is attributable to Uganda’s reduction in all-cause mortality and high fertility rate,^2^ but some growth is also due to in-migration.^9^ This rapid population growth may prevent ongoing open cohort studies like the RCCS from representatively sampling the growing target population.

Studies may address this challenge in two ways: they may prioritize longitudinal follow-up of existing residents by excluding new residents or they may scale down longitudinal follow-up efforts and prioritize representative sampling. For example, one demographic surveillance site in Burkina Faso prioritized enrolling new in-migrants over the longitudinal follow-up of out-migrants.^10^ In contrast, a cohort study in Malawi prioritized the follow-up of out-migrants instead of enrolling new in-migrants.^11^ In the case of the RCCS, rapid population growth and resource constraints led to the decision in 2008 to begin excluding residents of new households in newly built physical structures predominately comprised of in-migrating families.

Exclusion of new households from open population-based cohorts like the RCCS may bias estimates of community-level HIV seroprevalence if members of in-migrating households have higher or lower rates of HIV than local residents,^12^ (the latter because of a “healthy migrant effect”).^13–15^ Similarly, estimates of population-level rates of viremia among persons living with HIV may be biased by the exclusion of new households, and most evidence in SSA suggests worse HIV outcomes for migrants compared to non-migrants due to gaps in the continuity of care.^16–18^

Here, we explored how systematic exclusion of in-migrants in new structures may bias survey samples and alter HIV estimates. We first described characteristics of migrant individuals in new household structures, comparing them to both migrants and non-migrants in existing household structures. We then built an ensemble machine learning model to predict HIV seroprevalence and viremia in the excluded population.

## Methods

### The Rakai Community Cohort Study

The RCCS is an open population-based HIV surveillance cohort study in the Rakai region of South-Central, Uganda, established in 1994.^19^ At approximately 18 month intervals, the RCCS conducts a census of all residents, whether permanent or transient and regardless of age, in every household (old and new) within cohort community boundaries. The census collects sociodemographic data on each household member, including age, gender, marital status, residence status, each member’s relationship to the household head, and household assets.

Approximately two weeks after the census, the RCCS administers a survey interview to each consenting resident aged 15-49 years residing in old households. The survey collects sociodemographic, behavioural, health, and HIV service utilization data and a blood sample for the assessment of HIV prevalence, incidence, and HIV viremia among persons with HIV. Approximately 95% of eligible persons present in the household agree to both the interview and the blood draw.^20^ All participants are offered HIV results and post-test counselling on site.

For this analysis, we used data from 33 inland rural and peri-urban communities surveyed in RCCS round 19, conducted between June 2018 and November 2020.

### Migration and household structure classification

The RCCS defines a household as “an individual or group of individuals who eat their primary meals together and live together.”^20^ A household “structure” refers to the physical building in which members of the household reside. During each census, the RCCS identifies individuals living in newly identified structures who are members of entirely new households not previously censused in the RCCS. Since 2008, these individuals are included in the RCCS household census but are not eligible to participate in the survey.

Based on the household census, residents in the RCCS surveillance area were categorized according to whether they resided in a new or old physical structure and whether they were members of a newly established or old household. Old structures and old households were defined as being recorded on the census since any year prior to 2008. New structures and new households were defined as being recorded for the first time in year 2008 or after. Using new and old categorizations for both household and structure, the four categories of residents in the RCCS surveillance area are as follows (Figure 1):

**Figure 1.**
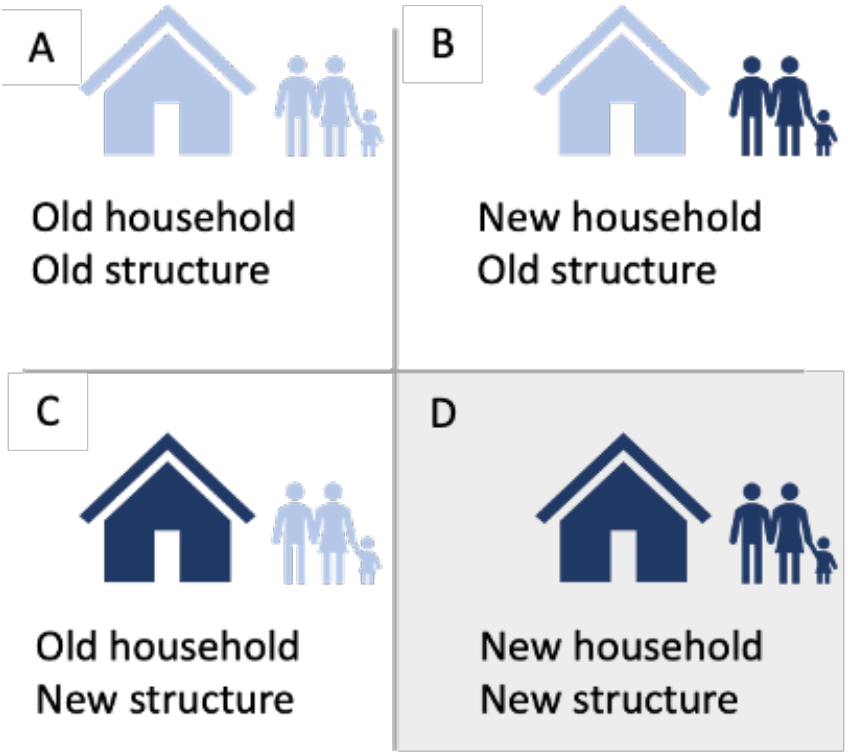
Four categories of household residents in the Rakai Community Cohort Study, by new or old household and new or old physical structure. Figure 1D is shaded grey because these residents were not eligible to participate in the survey.

A. Residents of old households in old structures: household censused prior to 2008 and members eligible to participate in the survey.
B. Residents of new households in old structures: household censused in 2008 or after and members eligible to participate in the survey.
C. Residents of old households in new structures: household censused prior to 2008 and members eligible to participate in the survey.
D. Residents of new households in new structures: household censused in 2008 or after and members not eligible to participate in the survey.

Since residents of new households in new structures (herein referred to as “new household residents” or “residents of new households”) have been excluded since 2008, we sought to compare them with residents of old structures (herein referred to as “old household residents” or “residents of old households”).

Residents of old households in new structures (Figure 1C) were excluded from the analysis because only 339 (56%) of the 601 residents had participated in the survey, comprising 1% of the analytic sample. These residents were similar to other residents of old households with regards to seroprevalence and viremia, so their exclusion was not expected to influence the results.

### Primary outcomes

Primary outcomes included population HIV seroprevalence and viremia. HIV seropositivity was assessed using a validated three rapid test algorithm.^21^ HIV viremia was defined as the proportion of the total population that had an HIV viral load of 1,000 copies/mL or more. We assigned a viral load of zero to all participants who tested HIV seronegative.

### Predictors

Census data were used to assess potential predictors of HIV seroprevalence and viremia. Individual predictors included gender, age group, marital status, relationship to the household head, and residence status (permanent or transient). Household-level predictors included region of residence, socio-economic status (SES), land ownership, household size, gender of the household head, and youth dependency ratio. SES was measured as previously described.^20^ Youth dependency ratio was defined as the number of household members under age 18 divided by the number of members aged 18 and above.^22^

### Descriptive analysis

We first measured the cumulative number of residents in new households and the cumulative number of new households over time. We then characterized residents in new and old households using the predictors described above, with differences assessed using chi-square tests. Analyses were additionally stratified by age (15-24 and 25-49) and by round in which new household was identified on the census (pre-2018 census or during the 2018 census) (reported in Supplement, Tables 1-2). Among residents in old households who participated in the survey, each predictor was assessed for its bivariate association with each outcome using a chi-square test of significance (reported in Supplement, Table 3).

To assess whether in-migrants in new households were different from in-migrants in old households, the analysis was repeated among in-migrants only. New in-migrants were individuals who have moved into the RCCS study area between the last survey round (August 2016 - May 2018) and this survey round (May 2018 - June 2020). Implicitly, all residents in new households were new in-migrants.

### Prediction model

In order to predict seroprevalence and viremia among residents in new households, an ensemble prediction model was built in Super Learner for each outcome. Super Learner uses cross validation to evaluate and weight the performance of multiple machine learning algorithms to build one ensemble model, thus maximizing efficiency while minimizing predictive error.^23^ In the first step of 10-fold cross-validation, each Super Learner weighted seven candidate algorithms (gam, biglasso, xgboost, glm.interaction, glm, bayesglm, and ranger) according to their prediction performance defined by the area under the receiver-operator-characteristic curve (AUC) (weights and descriptions of each algorithm are in the Supplement, Table 5). Another 10-fold cross-validation was then used to evaluate the performance of the overall ensemble model. Individuals living with the same household ID were grouped in each ensemble model to account for clustering of observations.

The model was used to predict seroprevalence and viremia probabilities for all residents in new households, and then for men, women, young people 15-24, and adults 25-49. The predicted prevalence in each of these groups was compared to the empirical prevalence in the observed sample. The predicted prevalence among residents of new households was multiplied by the size of the group to obtain the predicted number of residents who were HIV positive or viremic.

These numbers were combined with the number HIV positive or viremic in the observed sample to calculate the total seroprevalence and viremia prevalence for residents in both new and old households.

To account for uncertainty in each individual’s probability assignment for being HIV positive or viremic, everyone’s unknown final outcome was determined by simulating a Bernoulli trial with probability corresponding to the predicted probability of the outcome from the model. To account for uncertainty due to both sampling and individual level outcome assignment, the analysis was repeated on 100 bootstrapped samples of residents in new households.

## Results

There were 24,729 census-eligible adults, of whom 2,920 (12%) were new household residents and were thus excluded from the RCCS survey (Figure 2). 21,208 (86%) of censused individuals were old household residents, of whom 17,339 (82%) were eligible to participate in the survey and 11,942 (69%) were both eligible and present at the time of the survey. Residents who were not eligible to participate were either minors without parental consent, incapacitated, or had already been seen in another household. All residents in new households were new in-migrants compared to 20% (4,139) of those in old households.

**Figure 2.**
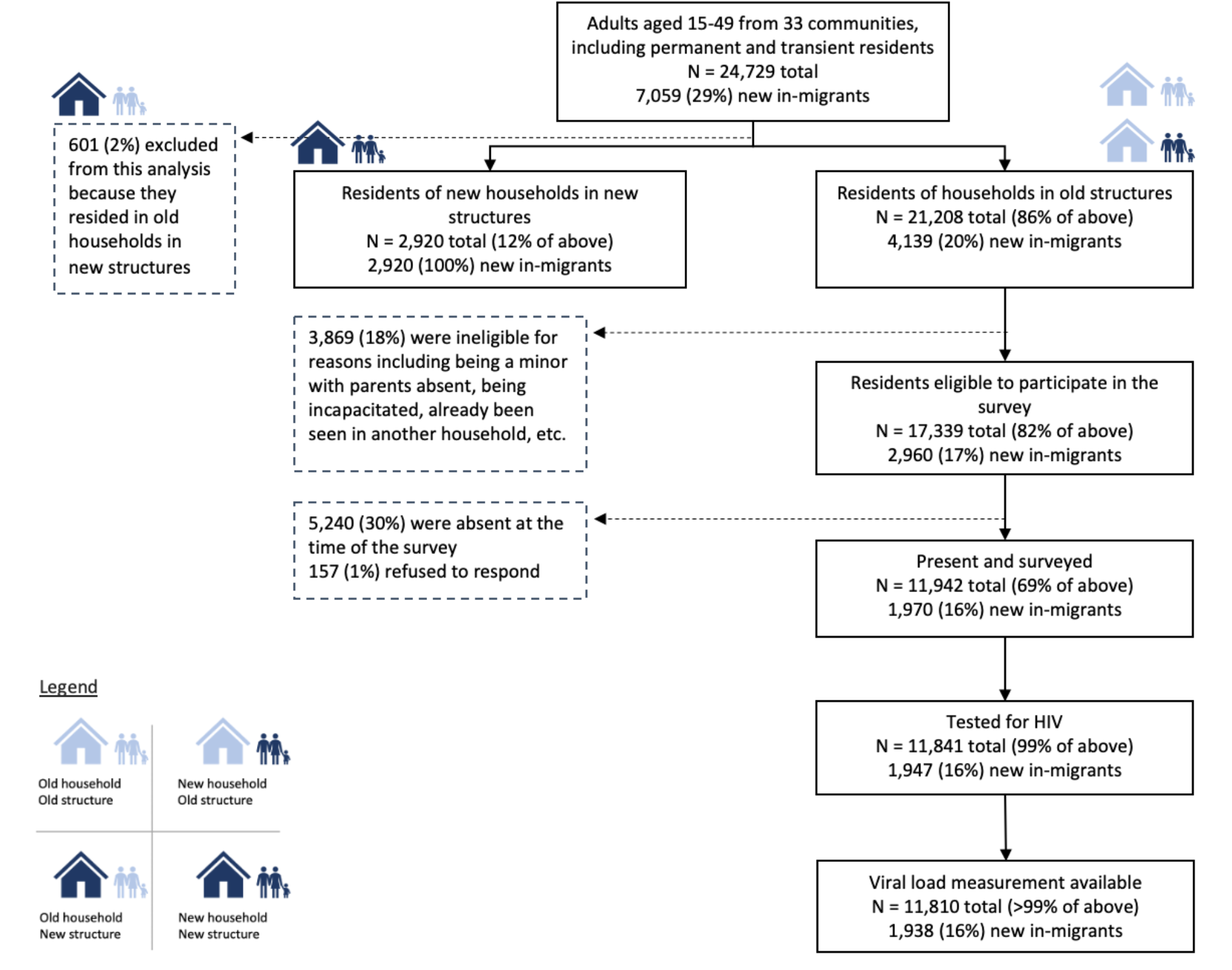
Study flow including number of participants.

Since 2009, the cumulative number of residents in new households increased from 90 to 2,920, with the steepest increase occurring between 2017 and 2020 (Figure 3). As of 2019 (survey round 19), 2,108/12,644 (17%) households and 2,920/24,729 (12%) residents were excluded from the RCCS survey.

**Figure 3.**
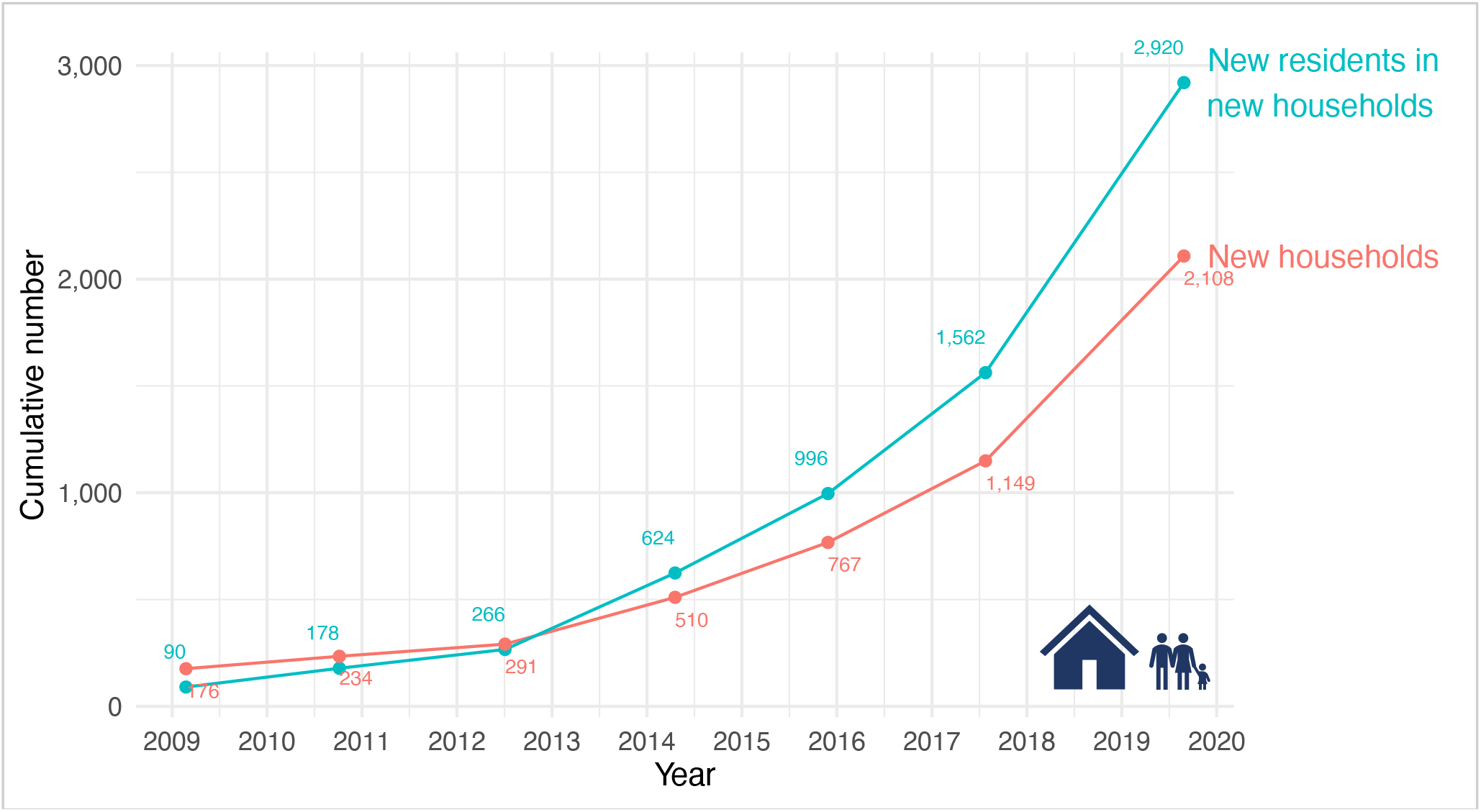
Cumulative number of residents and new households in new structures, by year in which new structure was identified on the census, 2008 – 2018, Rakai, Uganda.

When comparing residents in new households to residents in old households, the two groups differed with regards to all variables except for gender (Table 1). On average, residents in new households were more likely than those in old households to be middle-aged, married, the head of household, middle-SES, renters vs. owners, and from small households with less youth dependents.

**Table 1.**
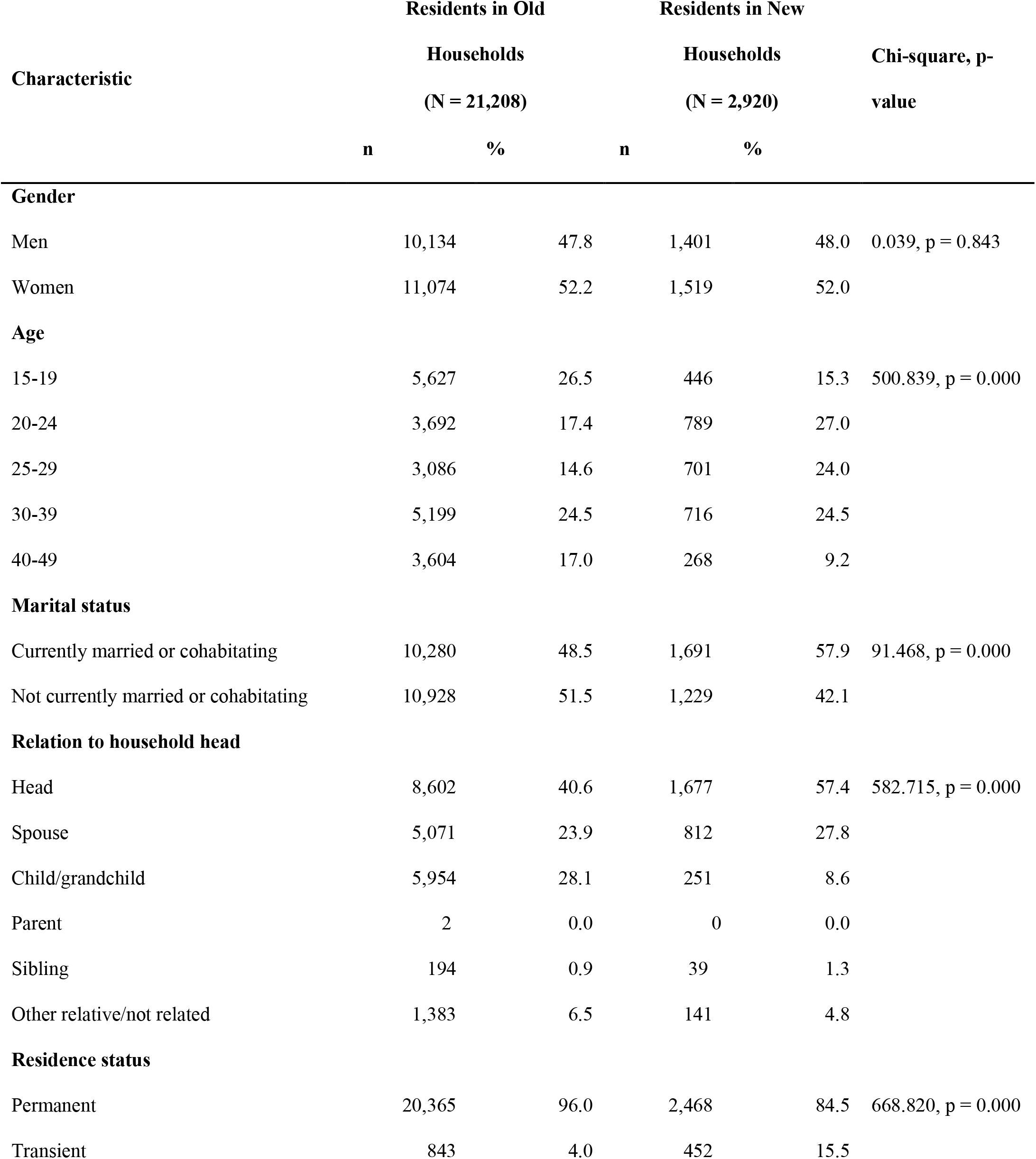
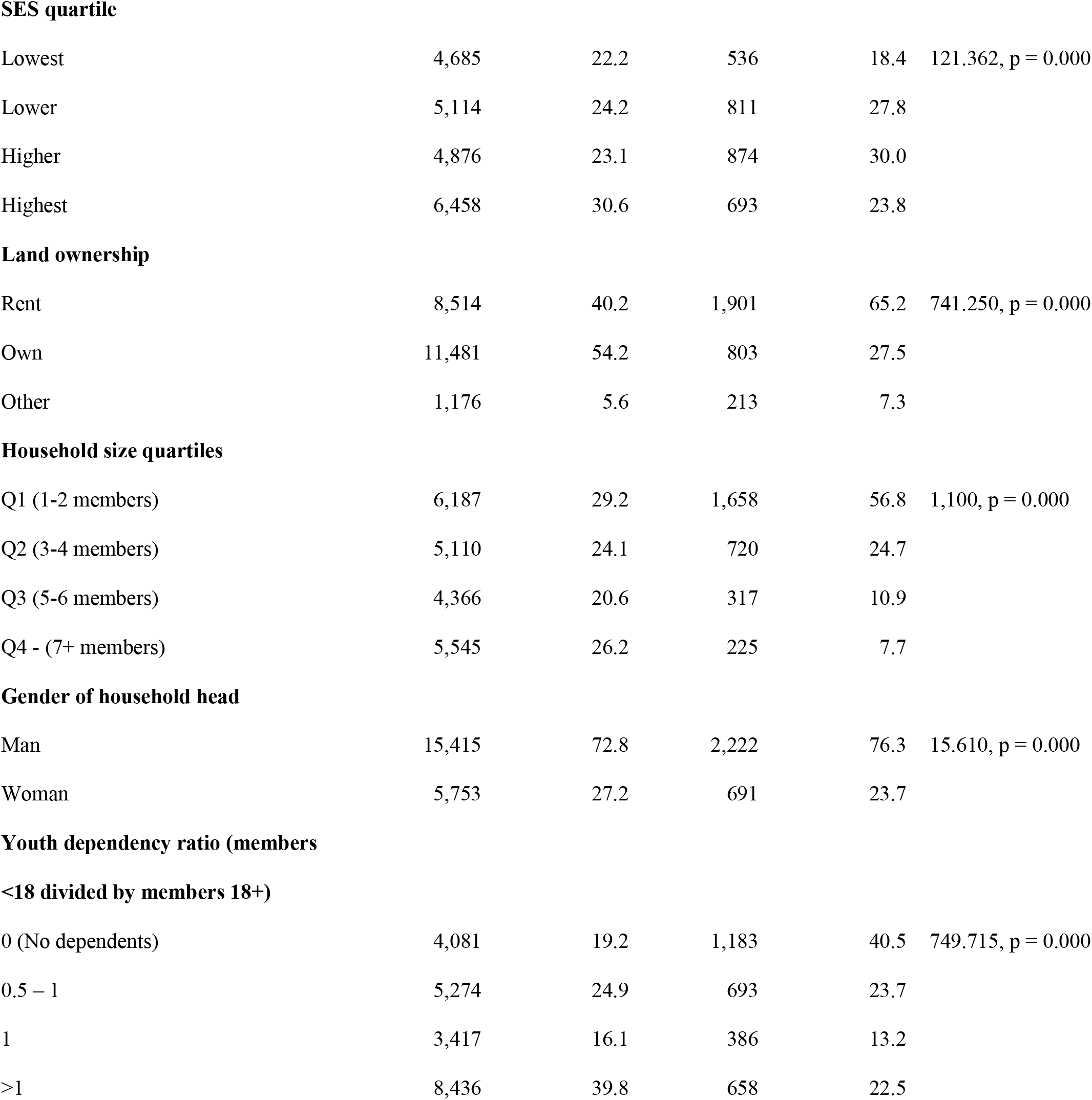
Characteristics of residents living in old and new households in round 19 of the Rakai Community Cohort, 2018 census (N = 24,128)

| Characteristic | Residents in Old |  | Residents in New |  | Chi-square, p-value |
| --- | --- | --- | --- | --- | --- |
|  | Households |  | Households |  |  |
|  | (N = 21,208) |  | (N = 2,920) |  |  |
|  | n | % | n | % |  |
| Gender |  |  |  |  |  |
| Men | 10,134 | 47.8 | 1,401 | 48.0 | 0.039, p = 0.843 |
| Women | 11,074 | 52.2 | 1,519 | 52.0 |  |
| Age |  |  |  |  |  |
| 15-19 | 5,627 | 26.5 | 446 | 15.3 | 500.839, p = 0.000 |
| 20-24 | 3,692 | 17.4 | 789 | 27.0 |  |
| 25-29 | 3,086 | 14.6 | 701 | 24.0 |  |
| 30-39 | 5,199 | 24.5 | 716 | 24.5 |  |
| 40-49 | 3,604 | 17.0 | 268 | 9.2 |  |
| Marital status |  |  |  |  |  |
| Currently married or cohabitating | 10,280 | 48.5 | 1,691 | 57.9 | 91.468, p = 0.000 |
| Not currently married or cohabitating | 10,928 | 51.5 | 1,229 | 42.1 |  |
| Relation to household head |  |  |  |  |  |
| Head | 8,602 | 40.6 | 1,677 | 57.4 | 582.715, p = 0.000 |
| Spouse | 5,071 | 23.9 | 812 | 27.8 |  |
| Child/grandchild | 5,954 | 28.1 | 251 | 8.6 |  |
| Parent | 2 | 0.0 | 0 | 0.0 |  |
| Sibling | 194 | 0.9 | 39 | 1.3 |  |
| Other relative/not related | 1,383 | 6.5 | 141 | 4.8 |  |
| Residence status |  |  |  |  |  |
| Permanent | 20,365 | 96.0 | 2,468 | 84.5 | 668.820, p = 0.000 |
| Transient | 843 | 4.0 | 452 | 15.5 |  |
| **SES quartile** |  |  |  |  |  |
| Lowest | 4,685 | 22.2 | 536 | 18.4 | 121.362, p = 0.000 |
| Lower | 5,114 | 24.2 | 811 | 27.8 |  |
| Higher | 4,876 | 23.1 | 874 | 30.0 |  |
| Highest | 6,458 | 30.6 | 693 | 23.8 |  |
| **Land ownership** |  |  |  |  |  |
| Rent | 8,514 | 40.2 | 1,901 | 65.2 | 741.250, p = 0.000 |
| Own | 11,481 | 54.2 | 803 | 27.5 |  |
| Other | 1,176 | 5.6 | 213 | 7.3 |  |
| **Household size quartiles** |  |  |  |  |  |
| Q1 (1-2 members) | 6,187 | 29.2 | 1,658 | 56.8 | 1,100, p = 0.000 |
| Q2 (3-4 members) | 5,110 | 24.1 | 720 | 24.7 |  |
| Q3 (5-6 members) | 4,366 | 20.6 | 317 | 10.9 |  |
| Q4 - (7+ members) | 5,545 | 26.2 | 225 | 7.7 |  |
| **Gender of household head** |  |  |  |  |  |
| Man | 15,415 | 72.8 | 2,222 | 76.3 | 15.610, p = 0.000 |
| Woman | 5,753 | 27.2 | 691 | 23.7 |  |
| **Youth dependency ratio (members****<18 divided by members 18+)** |  |  |  |  |  |
| 0 (No dependents) | 4,081 | 19.2 | 1,183 | 40.5 | 749.715, p = 0.000 |
| 0.5 – 1 | 5,274 | 24.9 | 693 | 23.7 |  |
| 1 | 3,417 | 16.1 | 386 | 13.2 |  |
| >1 | 8,436 | 39.8 | 658 | 22.5 |  |

When comparing new in-migrants in new households to new in-migrants in old households, the two groups differed with regards to all variables including gender (Table 2). Migrants in new households were less likely to be women (52.0%) compared to migrants in old households (58.7%). Compared to those in old households, the excluded population of new in-migrants were more likely to be men, aged 25-39, married, the household head, higher SES, owners vs. renters, and of household with a man as household head.

**Table 2.**
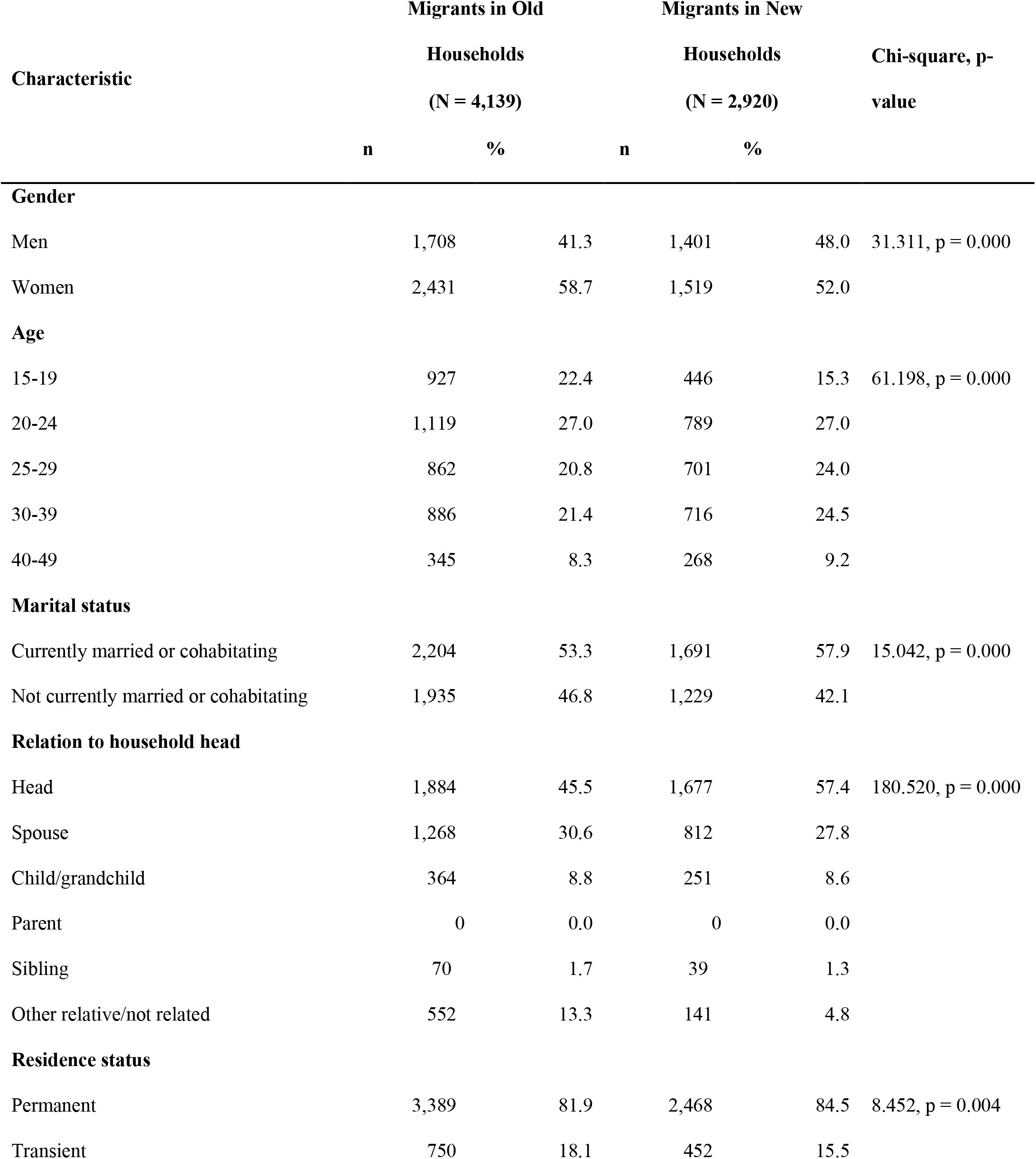
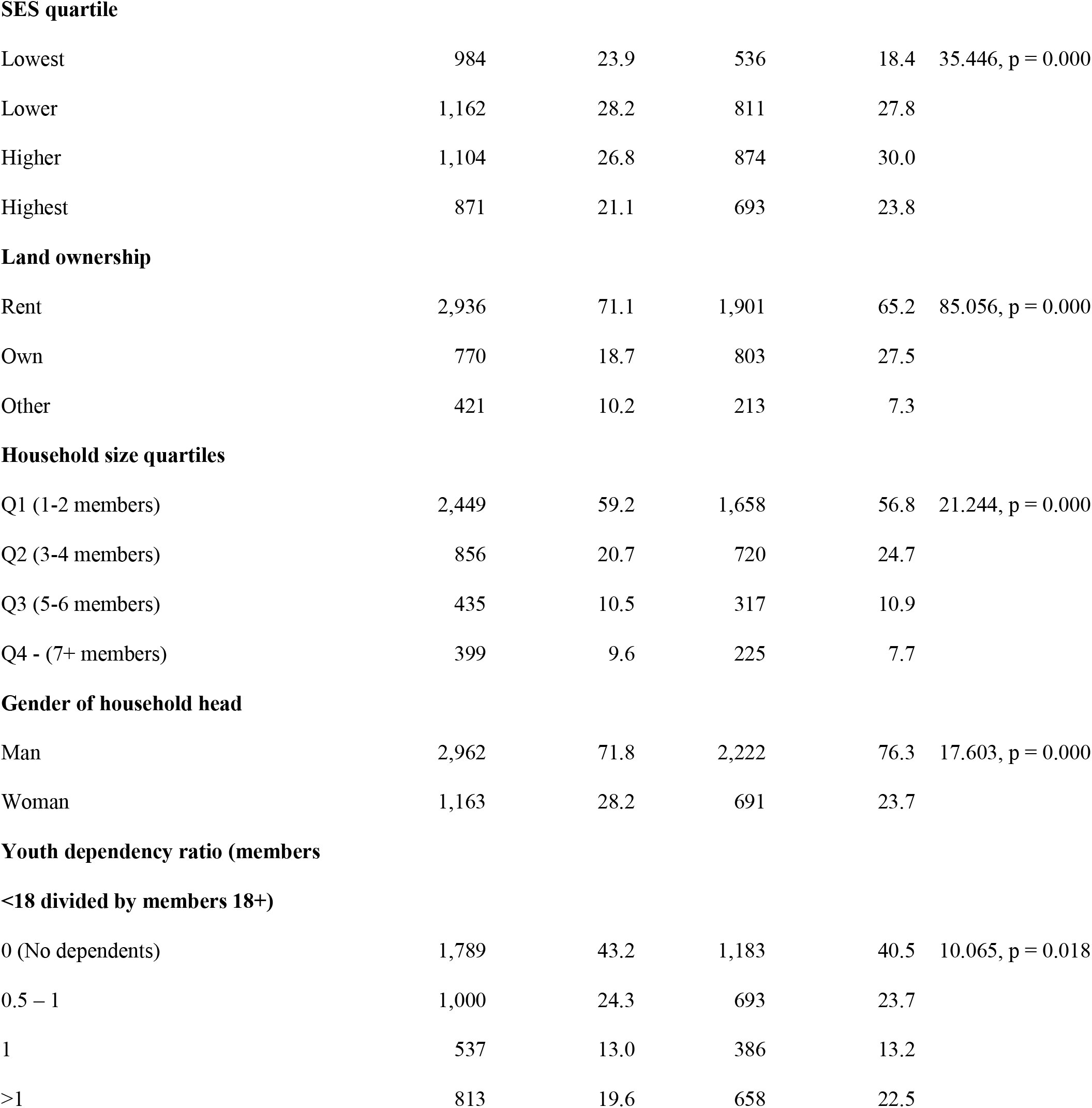
Characteristics of new in-migrants living in old and new households in round 19 of the Rakai Community Cohort, 2018 census (N = 7,059)

| Characteristic | Migrants in Old |  | Migrants in New |  | Chi-square, p-value |
| --- | --- | --- | --- | --- | --- |
|  | Households |  | Households |  |  |
|  | (N = 4,139) |  | (N = 2,920) |  |  |
|  | n | % | n | % |  |
| Gender |  |  |  |  |  |
| Men | 1,708 | 41.3 | 1,401 | 48.0 | 31.311, p = 0.000 |
| Women | 2,431 | 58.7 | 1,519 | 52.0 |  |
| Age |  |  |  |  |  |
| 15-19 | 927 | 22.4 | 446 | 15.3 | 61.198, p = 0.000 |
| 20-24 | 1,119 | 27.0 | 789 | 27.0 |  |
| 25-29 | 862 | 20.8 | 701 | 24.0 |  |
| 30-39 | 886 | 21.4 | 716 | 24.5 |  |
| 40-49 | 345 | 8.3 | 268 | 9.2 |  |
| Marital status |  |  |  |  |  |
| Currently married or cohabitating | 2,204 | 53.3 | 1,691 | 57.9 | 15.042, p = 0.000 |
| Not currently married or cohabitating | 1,935 | 46.8 | 1,229 | 42.1 |  |
| Relation to household head |  |  |  |  |  |
| Head | 1,884 | 45.5 | 1,677 | 57.4 | 180.520, p = 0.000 |
| Spouse | 1,268 | 30.6 | 812 | 27.8 |  |
| Child/grandchild | 364 | 8.8 | 251 | 8.6 |  |
| Parent | 0 | 0.0 | 0 | 0.0 |  |
| Sibling | 70 | 1.7 | 39 | 1.3 |  |
| Other relative/not related | 552 | 13.3 | 141 | 4.8 |  |
| Residence status |  |  |  |  |  |
| Permanent | 3,389 | 81.9 | 2,468 | 84.5 | 8.452, p = 0.004 |
| Transient | 750 | 18.1 | 452 | 15.5 |  |
| **SES quartile** |  |  |  |  |  |
| Lowest | 984 | 23.9 | 536 | 18.4 | 35.446, p = 0.000 |
| Lower | 1,162 | 28.2 | 811 | 27.8 |  |
| Higher | 1,104 | 26.8 | 874 | 30.0 |  |
| Highest | 871 | 21.1 | 693 | 23.8 |  |
| **Land ownership** |  |  |  |  |  |
| Rent | 2,936 | 71.1 | 1,901 | 65.2 | 85.056, p = 0.000 |
| Own | 770 | 18.7 | 803 | 27.5 |  |
| Other | 421 | 10.2 | 213 | 7.3 |  |
| **Household size quartiles** |  |  |  |  |  |
| Q1 (1-2 members) | 2,449 | 59.2 | 1,658 | 56.8 | 21.244, p = 0.000 |
| Q2 (3-4 members) | 856 | 20.7 | 720 | 24.7 |  |
| Q3 (5-6 members) | 435 | 10.5 | 317 | 10.9 |  |
| Q4 - (7+ members) | 399 | 9.6 | 225 | 7.7 |  |
| **Gender of household head** |  |  |  |  |  |
| Man | 2,962 | 71.8 | 2,222 | 76.3 | 17.603, p = 0.000 |
| Woman | 1,163 | 28.2 | 691 | 23.7 |  |
| **Youth dependency ratio (members****<18 divided by members 18+)** |  |  |  |  |  |
| 0 (No dependents) | 1,789 | 43.2 | 1,183 | 40.5 | 10.065, p = 0.018 |
| 0.5 – 1 | 1,000 | 24.3 | 693 | 23.7 |  |
| 1 | 537 | 13.0 | 386 | 13.2 |  |
| >1 | 813 | 19.6 | 658 | 22.5 |  |

### Predicted HIV seroprevalence and viremia among residents in new households

#### HIV seroprevalence

The AUC of the seroprevalence prediction model was 0.78 (0.74, 0.80) (Supplement, Table 5). Overall, predicted seroprevalence for new household residents was 11.4% (95% Confidence Interval (CI): 10.2, 12.3%) compared to 11.8% in the observed sample (Figure 4). Inclusion of new household residents in the total estimate for seroprevalence did not significantly change seroprevalence estimates for the total population (11.7% [11.4, 11.9]). However, for young people aged 15-24, inclusion of new household residents, with a predicted seroprevalence of 5.1% (3.6, 6.1), resulted in higher total seroprevalence (3.1% [2.8, 3.34]) for both new and old household residents in that age strata compared to the observed sample (2.6%).

**Figure 4.**
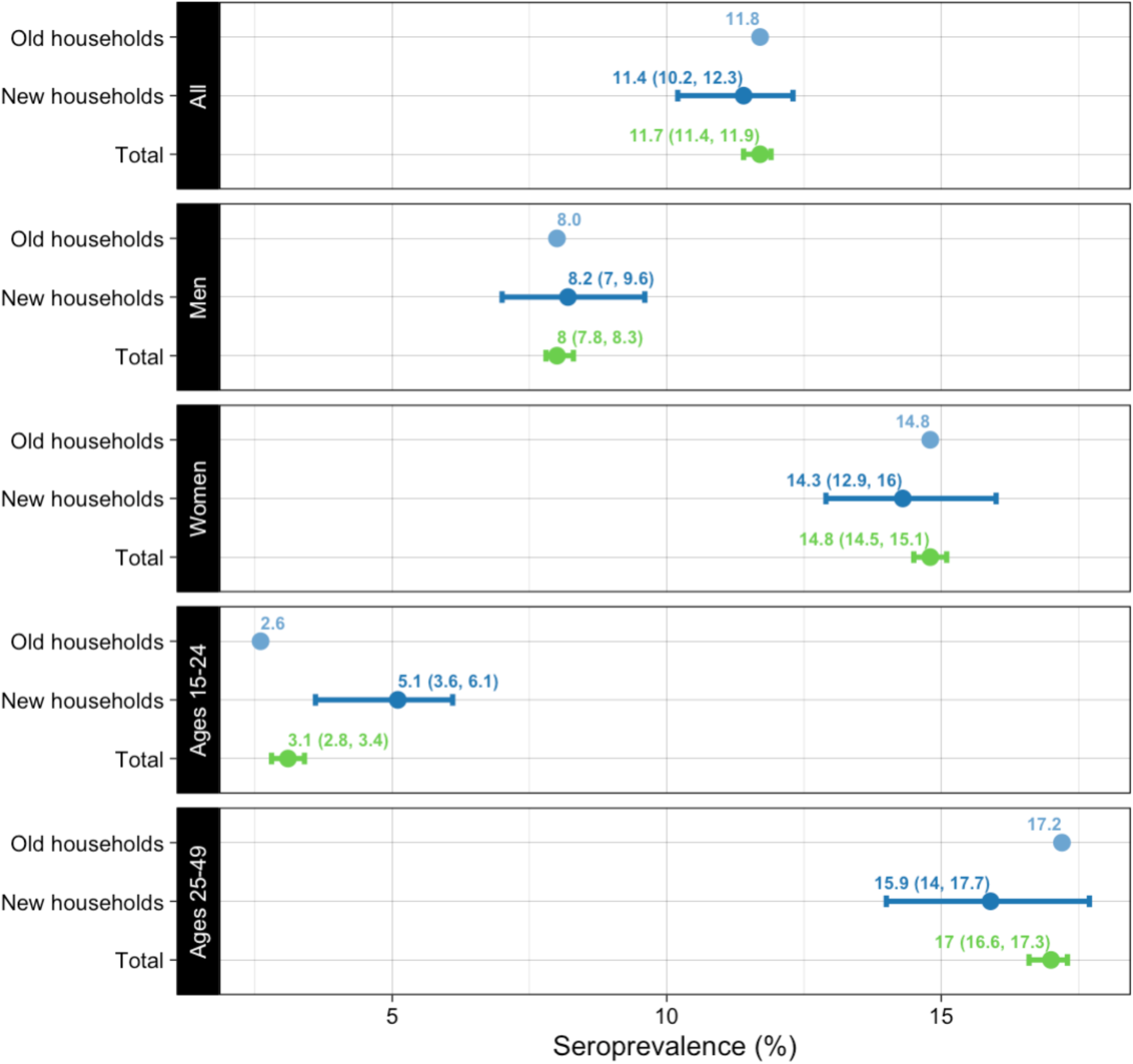
Seroprevalence in the RCCS Surveillance Sample, excluding and including predicted values for new households, Rakai, Uganda 2019-2020 (N=14,679)

#### Viremia

The AUC for the viremia prediction model was low relative to that of the seroprevalence prediction model at 0.69 (0.58, 0.83) (Supplement, Table 5). Inclusion of new household residents, with a predicted viremia prevalence of 2.78% (2.2, 3.3), resulted in a 12% higher total viremia prevalence (1.9% [1.8, 2.0]) for both new and old household residents compared to the observed sample (1.7%) (Figure 5). Similar findings were observed for all sub-groups by age and gender, with the comparatively highest viremia estimate predicted for young people aged 15-24 (1.1% [0.9, 1.2]) in the total population compared to viremia prevalence of 0.8% among residents in old households.

**Figure 5.**
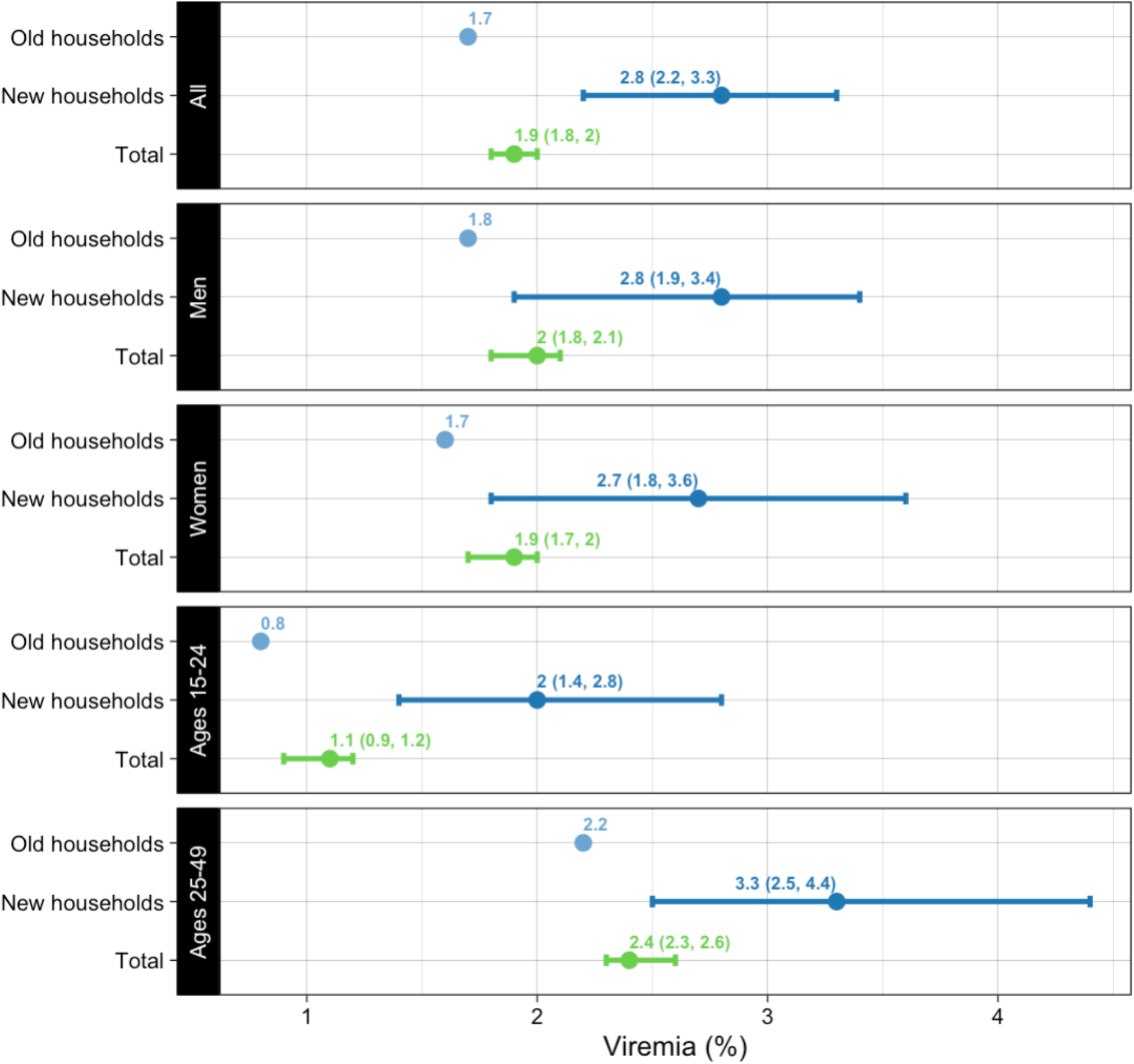
Viremia in the RCCS Surveillance Sample, excluding and including predicted values for new households, Rakai, Uganda 2019-2020 (N=14,648)

## Discussion

Longitudinal population-based cohorts have contributed immensely to the HIV response in Africa, but rapid population growth poses significant challenges to open cohorts like the RCCS. While there were few new households in 2008 when they were first excluded, the population in new households has expanded with time, and now substantively differs from the included surveillance sample on various sociodemographic characteristics. These differences did not translate into significantly different HIV seroprevalence rates overall, but modest biases may exist in some age-specific seroprevalence estimates and viremia estimates may be biased across all sub-groups.

Considering that all residents in new households were recent in-migrants, biases in the age-specific seroprevalence estimates illustrate the heterogeneity of migrant groups. Different types of migrants have different levels of HIV risk. Among 15-24-year-olds, there may be a migrant health penalty for those arriving in new households because their seroprevalence was higher than the included sample. Young people in Rakai who migrate have been shown to engage in higher HIV risk behaviours compared to their non-migrant counterparts.^24^ These risky behaviours often coincide with the transition from school to work, when many mobile young people engage in risky occupations associated with HIV risk, like boda-boda drivers, truck drivers, and bar and restaurant workers.^25^ Among 25-49-year-olds, the opposite seems to be true: there may be a healthy migrant effect for those arriving in new households because their estimated seroprevalence was lower than the included sample. Since many adults aged 25-49 in new households were married men and the head-of-household, it may be that this group of migrants travelled for family or greater economic opportunity and not due to circumstances that independently drive HIV risk.

Unlike seroprevalence, viremia rates are projected to be modestly but consistently higher across all sub-groups in new households, compared to old households, thereby increasing the rate of viremia in the total population. This provides evidence for the migrant health penalty, which posits that individuals who migrate experience intrinsic risks that heighten their risk for poor HIV outcomes.^16^ Mobile populations may face structural barriers to care that lead to treatment interruptions,^17^ which increases risk of viremia and other poor treatment outcomes.^26,27^ In Namibia, high viremia rates were found along transport corridors, signifying that high rates of viremia among migrants may also correspond to high community-level viremia.

Furthermore, excluding certain migrants from surveillance programs could mean that the wrong inferences are made about the health state and risks for migrant populations. For example, cohorts that only include permanent migrants with stronger social ties might produce evidence for the migrant experience that does not represent more transient or more marginalized migrants. In the case of the RCCS, the cohort may underrepresent migrant men, given that migrant men were more likely to be living in new household structures than old household structures. Since migrant men are more likely to be viremic,^28^ it is possible that viremia among migrants in the RCCS is higher than would be estimated with current data.

Our study had several limitations. First, the prediction model performance was moderate for seroprevalence and relatively weaker for viremia, which has been observed in other HIV ensemble modelling studies.^29^ This may be due to the relative lack of data on new residents: for example, sexual behaviours like age-disparate sexual partnerships predict seroprevalence,^30^ and health behaviours like alcohol use predict viremia,^31^ and these data were not available for residents of new households. Nonetheless, many of the included predictors have been demonstrated to reliably predict HIV outcomes,^28,32,33^ and a sub-analysis of RCCS survey data confirmed this (Supplement, Table 1). Second, this analysis assumes that the risk factors for HIV seroprevalence and viremia are the same for residents of new households as for residents of old households, but this assumption is difficult to prove. Finally, it is possible that we underestimated or overestimated seroprevalence or viremia among residents in new households because we do not know who would have been present and survey-eligible had the household been included in the survey. In the survey sample, residents in old households are sometimes excluded from the survey because they have died, are incapacitated at the time of the survey, or refuse to respond. If HIV outcomes differ for eligible and ineligible participants, then this could partially explain results.

While we did not find substantial potential biases from excluding new household residents in the RCCS, our results show this population is growing and observed differences may become more problematic over time. The characteristics of in-migrants may change over time as well, so their exclusion could bias HIV estimates differently in the future. An increasing number of people are moving for climate change-related reasons like food security,^34^ and women make up an increasing proportion of internal migrants in Africa.^35–37^ Given the critical role of longitudinal population-based cohorts in African HIV surveillance, these studies should routinely monitor and be transparent about any potential biases introduced by demographic change and resulting modifications to study inclusion criteria. Lastly, results from studies that prioritize representativeness should be triangulated with those from studies that maintain longitudinal follow-up to improve overall understanding of the African HIV epidemic.

## Data Availability

All data produced in the present study are available upon reasonable request to the authors and to the Rakai Health Sciences Program.

https://www.rhsp.org/

